# Sex, age and HIV-1 status influence complement activation against SARS-CoV-2 mRNA vaccine Spike protein

**DOI:** 10.64898/2026.11.04.26366940

**Authors:** Dylan J. Tuttle, Phyllis C. Tien, Nicolas Sluis-Cremer, Robbie B. Mailliard, Charles R. Rinaldo, Ernesto T. A. Marques

## Abstract

Regulation of complement activity influences immunity and clinical outcomes of SARS-CoV-2 infections. Here we investigated antibody responses to SARS-CoV-2 vaccination and complement activity in men (age 36 to 86) and women (age 39 to 83) with (MWH, WWH) and without (MWOH, WWOH) HIV-1. Antibody responses in WWH were lower than in MWH. WWH also had negligible change in blood C3a post-immunization, whereas MWH had greater C3a levels. We investigated the levels of C5, C5a, C1-INH, factor H, and circulating immune complex-C1q (CIC-C1q) binding to understand the regulation of complement activity. We found that C1-INH and anti-S antibody levels were driving the differences in C3 convertase activity in MWH. No significant differences were found between C5 and C5a in both cohorts, suggesting that C5 convertase is not being activated. These results show that biological sex may impact SARS-CoV-2 immune responses and highlights the need for further investigation.

## Introduction

SARS-CoV-2 mRNA vaccines provide effective immune responses against SARS-CoV-2 infection in people with HIV (PWH) [1, 2]. However, PWH can have inadequate antibody response to SARS-CoV-2 vaccines compared to people without HIV-1 (PWOH). We previously found that men with HIV-1 (MWH) respond to SARS-CoV-2 mRNA vaccines with lower binding IgG and neutralizing antibodies, along with a higher baseline complement activity than men without HIV-1 (MWOH) [3]. It is unclear whether these findings can be generalized to women with HIV-1 (WWH).

Women mount greater antibody responses to vaccination than men when age is not considered [4, 5]. Unfortunately, there is a noticeable gap in the information on differences in the response to the SARS-CoV-2 mRNA vaccines between MWH and women with HIV (WWH). To address this issue, we investigated vaccine-induced antibodies and their effector functions in men compared to women of similar age with (MWH, WWH) and without (MWOH, WWOH) HIV.

Antibody-dependent complement activation is an important component of the response to SARS-CoV-2 mRNA vaccines [6]. However, complement overactivity has been associated with greater COVID-19 disease severity [7-10]. We previously documented that MWH have greater complement activation following vaccination than MWOH [3], however the antibody dependent complement activation in WWH and WWOH is not known. This study addresses this significant knowledge gap.

## Methods

### Participants and clinical samples

Serum and plasma samples were obtained by the University of California San Franscisco and University of Pittsburgh clinical research sites in the MACS – WIHS Combined Cohort Study (MWCCS). The characteristics of this study population have been previously described [11]. All included participants were vaccinated with a two-dose regimen of one of the mRNA vaccines against the Washington (WA) virus strain. All participants provided informed consent, and the study was approved by the University of California San Franscisco and University of Pittsburgh Institutional Review Boards.

### Enzyme-linked immunosorbent assay (ELISA)

Serum samples were evaluated for IgG binding to SARS-CoV-2 S1 (SinoBiological) and N (RayBiotech) proteins (WA strain) [12] using an in-house developed (research grade) indirect ELISA described previously and in the supplemental material [3]. The lower limit of quantitation for the ELISA was defined as three times the standard deviation of the negative control.

### Measurement of complement factors

Measurement of serum C3 and C3a levels was carried out in-house as described previously and in the supplemental material, with the lower limit of quantitation being 0.1 and 0.03ng/mL, respectively [12]. C1q-CIC was measured using an ELISA kit (QuidelOrtho) as per the manufacturer’s specifications.

### Complement activation assay

Complement activation was assessed using a bead-based flow cytometry assay as described previously and in the supplemental material [3]. The reported data represent an average mean fluorescence intensity of the 3 replicate experiments performed on each sample.

### Statistical analysis

Analyses (Prism software version 10; GraphPad) included use of nonparametric Mann-Whitney U tests and Kruskal-Wallis ANOVAs with Dunn *post hoc* test for continuous variables without pairing, Wilcoxon matched pairs tests for paired continuous variables, and Fisher exact tests for categorical variables. Spearman correlation was used to assess associations between two variables. Analyses were not corrected for confounders due to the small sample size. A p-value of less than or equal to 0.05 was considered statistically significant.

## Results

### Study population

The average age for WWH was 58.5 (range 39-83) and for WWOH was 51 (range 40-69), highlighting that the WWH group was an average of 7.5 years older than the WWOH group (p=0.008; Table 1). By contrast, MWH were on average 5 years younger, but the difference in mean age was not statistically different (MWH: 62 (36-86), MWOH: 67 (31-84); Table 1) [3]. WWH also had lower nadir CD4^+^ T cell counts than WWOH (P=0.0001; Table 1). MWH on average had CD4^+^ T cell counts within the healthy range (>500cells/μL) (Table 1) [3]. The distribution of the two vaccines, mRNA-1273 and BNT-162b2 that were administered to WWH and WWOH were similar (p=0.371), as was the SARS-CoV-2 seropositivity rate (p=0.799), which was also true for MWH and MWOH. To discern between vaccine-induced immune responses from those elicited from natural exposure to SARS-CoV-2, participants were assessed for anti-nucleoprotein (anti-N) IgG and excluded from the study if tested positive.

**Table 1.**
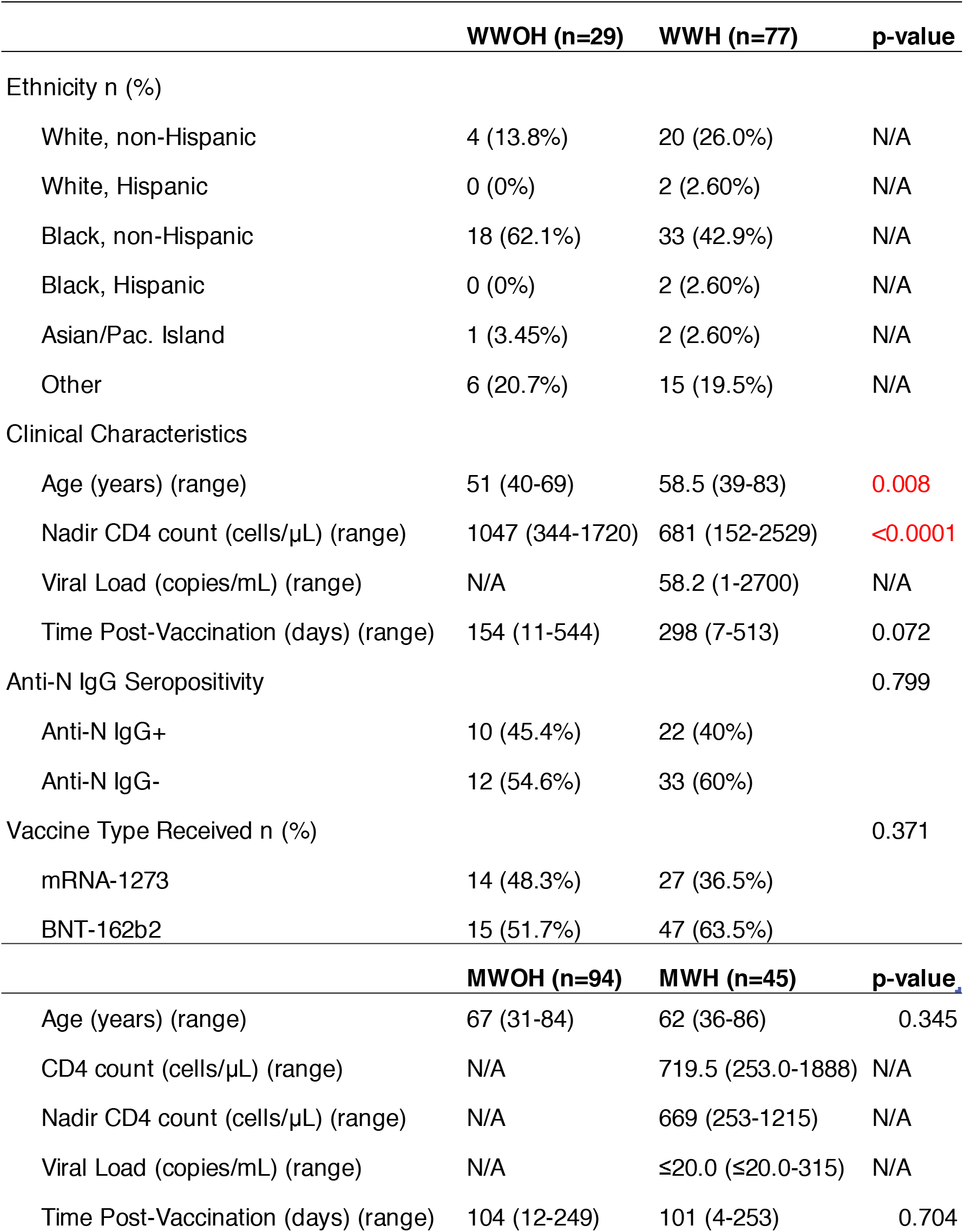
Demographic characteristics of WWOH and WWH from the UCSF Clinical Research Site. All participants were on ART at the time of sample collection. Samples were collected on a rolling basis throughout 2021. Data are represented as number (%) or median (range). Two-tailed Fisher Exact and Mann-Whitney U tests were used to compare between the groups. Time post-vaccination was defined as the time between the second dose of the 2-dose immunization regimen and sample collection. P-values in red denote statistical significance. Abbreviations: IgG, immunoglobulin G; WWH, women with HIV-1; WWOH, women without HIV-1; N, nucleocapsid.

### Antibody titers and overall baseline complement activity

To understand vaccine-induced immune responses in WWH versus MWH, we measured anti-spike 1 (anti-S1) IgG titers. We found that MWH had significantly higher levels of anti-S1 IgG titers compared to WWH (p<0.0001; Figure 1), and in MWOH compared to WWH (p<0.0001; Figure 1). Interestingly, approximately half of all included women had undetectable levels of anti-S1 IgG antibody, and the proportion was not different between WWH and WWOH (p=0.719; data not shown). Interestingly, the proportion of vaccine non-responders did not associate with age, and women between 40 and 80 years old did not have detectable anti-S1 IgG responses (Figure S1). In contrast, there was a lower proportion of non-responders in MWH, suggesting that WWH generated lower humoral responses than MWH.

**Figure 1.**
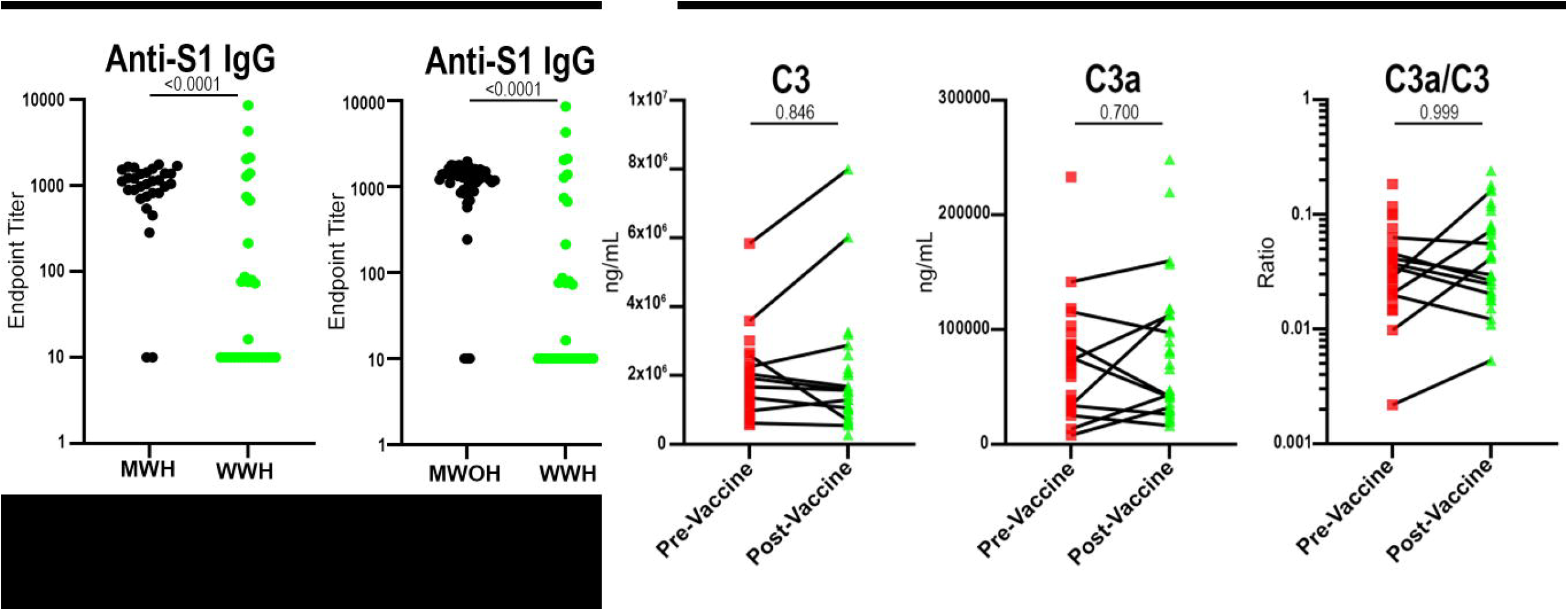
Antibody and complement levels in WWH. Anti-S1 IgG levels in WWH (n=32), MWH (n=30) and MWOH (N=52) were measured by indirect ELISA. C3 and C3a were determined in WWH (n=10) pre- and post-vaccine using capture ELISA, and the C3a/C3 ratio was determined by taking the C3a levels and dividing by the C3 levels at each timepoint. Participants are anti-N IgG negative, and two-dose vaccinated with a SARS-CoV-2 mRNA vaccine. Mann-Whitney U tests were used to compare between participant groups, and Wilcoxon matched pairs tests were used to compare between pre- and post-vaccine timepoints. A p-value of 0.05 is considered statistically significant. Abbreviations: IgG, immunoglobulin G; WWH, women with HIV-1; WWOH, women without HIV-1; N, nucleocapsid.

We previously found that MWH have greater complement activation following immunization than MWOH through the release of C3a post-vaccination [3]. Measurement of C3a, which is a chemoattractant for immune cells, is therefore useful in understanding the consumption of C3. When comparing levels of C3 and C3a in WWH pre-vaccine and post-vaccine, we did not find a significant change in serum levels (p=0.896 and 0.700, respectively; Figure 1), and consequently the C3a/C3 ratio between pre-and post-vaccine timepoints remained unchanged (p=0.999; Figure 1). Overall, WWH had a comparable response to the vaccines as WWOH.

We next measured regulatory factors within the alternative and classical complement cascades, as both cascades share the C3 convertase amplification loop but have different compositions of these complexes, and thus, different regulators. Factor H is one of the regulators unique to the alternative pathway, which destabilizes the C3 convertase. Here we found no difference in factor H levels at the post-vaccine timepoint between MWH and MWOH (p=0.396; Figure 2A). However, there was a decrease in C1-inhibitor (C1-INH), which regulates the formation of the C3 convertase in the classical pathway through C1q, in both MWOH and MWH post-vaccine (p=0.003 for both; Figure 2B). MWOH and MWH also had a decline in C1q-circulating immune complexes (CIC-C1q) post-vaccination (p=0.029 and 0.001, respectively; Figure 2B).

**Figure 2.**
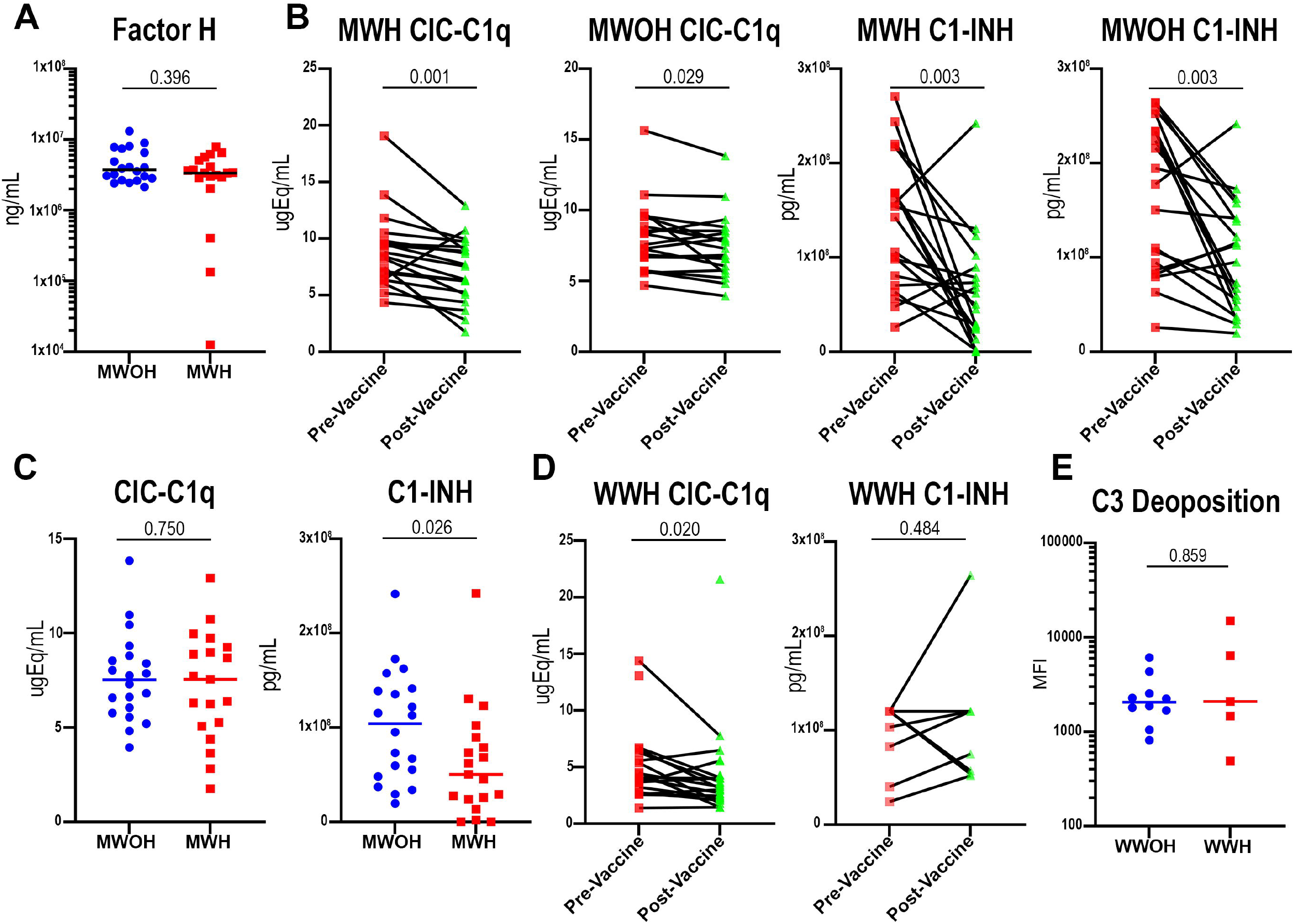
Complement regulators in MWH, MWOH and WWH. (A). Factor H was measured in MWH (n=19) and MWOH (n=20) using capture ELISA. (B). CIC-C1q and C1-INH measurements were determined in MWH (n=19) and MWOH (n=19) pre- and post-vaccine using a capture ELISA kit. (C). CIC-C1q and C1-INH measurements were determined in MWH (n=19) and MWOH (n=19) using a capture ELISA kit. (D). CIC-C1q (n=17) and C1-INH (n=10) were measured in WWH pre- and post-vaccine using capture ELISA. (E). C3 deposition in WWH (n=10) and WWOH (n=5) was determined via bead-based flow cytometry assay. Participants are anti-N IgG negative except for the CIC-C1q WWH, and two-dose vaccinated with a SARS-CoV-2 mRNA vaccine. Mann-Whitney U tests were used to compare between participant groups, and Wilcoxon matched pairs tests were used to compare between pre- and post-vaccine timepoints. A p-value of 0.05 is considered statistically significant. Abbreviations: IgG, immunoglobulin G; MWH, men with HIV-1; MWOH, men without HIV-1; WWH, women with HIV-1; WWOH, women without HIV-1; N, nucleocapsid; CIC-C1q, circulating immune complexes-C1q; C1-INH, C1 inhibitor.

There was no difference in CIC-C1q levels between MWH and MWOH (p=0.750; Figure 2C). However, MWH did have significantly lower levels of C1-INH post-vaccine compared to MWOH (p=0.026; Figure 2C). We observed that WWH had lower CIC-C1q levels post-vaccine (p=0.020; Figure 2D), but that there was no change in the C1-INH (p=0.484; Figure 2D). Furthermore, neither WWH or WWOH displayed significant differences in C3 deposition (p=0.859; Figure 2E).

### Complement activity terminates following C3 deposition in MWH and is absent in WWH

To understand the extent to which the complement cascade proceeded, we measured serum C5a levels in MWH and MWOH pre-and post-vaccine and found no significant differences (p=0.891 and 0.465, respectively; Figure 3A). There were also no differences in serum C5a levels at the post-vaccine timepoint between MWH and MWOH (p=0.751; Figure 3B). Like MWH, there was no significant change in C5a levels in WWH (p=0.275; Figure 3C).

**Figure 3.**
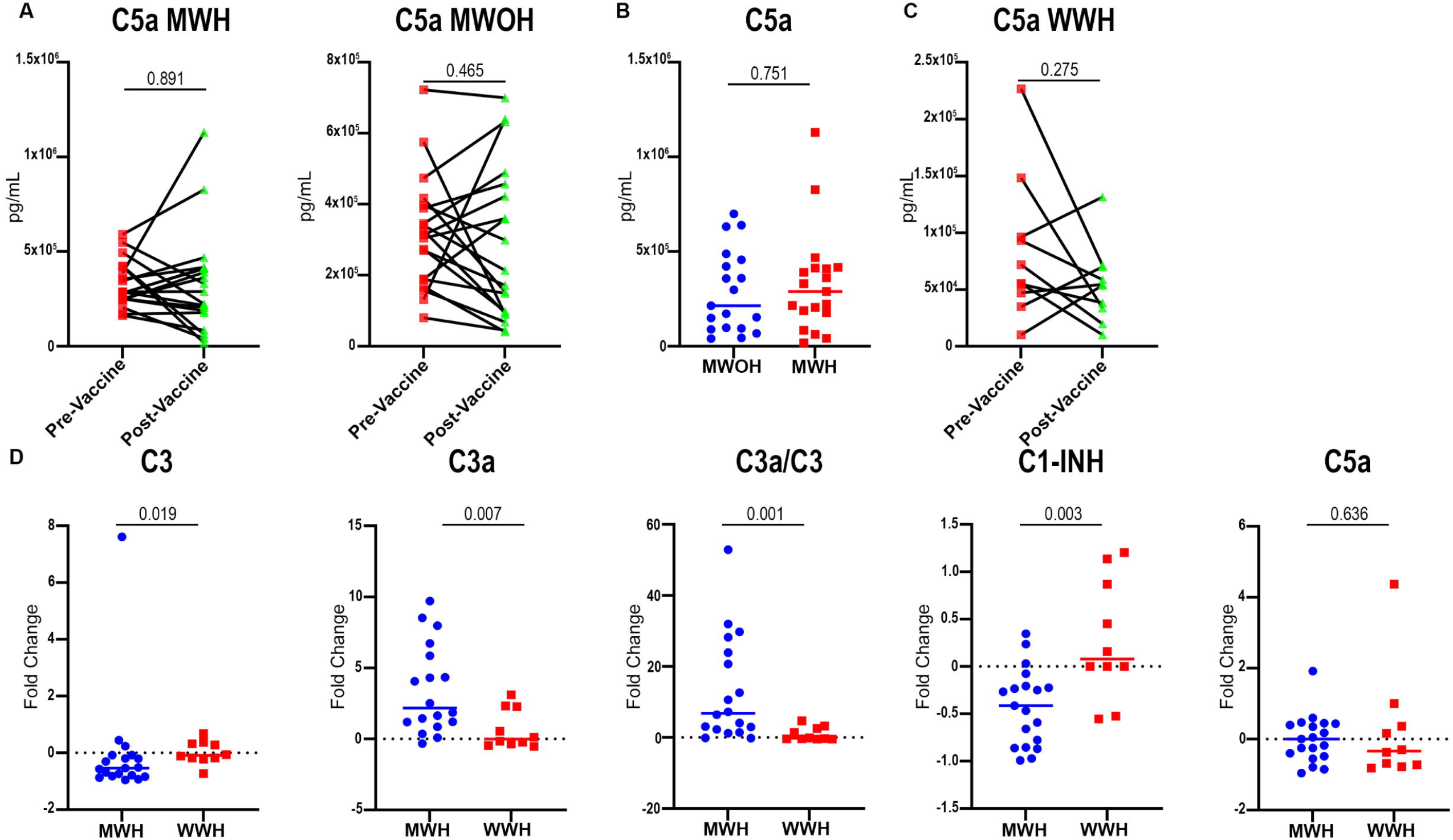
Complement activity in MWH and WWH. (A-C). C5a levels were determined pre- and post-vaccine in MWH (n-19), MWOH (n=19) and WWH (n=10) using capture ELISA. (D). Fold change in complement factors in MWH (n=19) and WWH (n=10) were determined by taking the difference of the post-vaccine level and pre-vaccine level, then dividing by the pre-vaccine level. Participants are anti-N IgG negative, and two-dose vaccinated with a SARS-CoV-2 mRNA vaccine. Mann-Whitney U tests were used to compare between participant groups, and Wilcoxon matched pairs tests were used to compare between pre- and post-vaccine timepoints. A p-value of 0.05 is considered statistically significant. Abbreviations: MFI, mean fluorescence intensity; MWH, men with HIV-1; MWOH, men without HIV-1; WWH, women with HIV-1; N, nucleocapsid; CIC-C1q, circulating immune complexes-C1q; C1-INH, C1 inhibitor.

Finally, we compared complement activity directly between men and women based on the fold-change of each of the serum complement factors we measured from pre- to post-vaccine. We found that MWH had significantly higher levels of C3 (p=0.019; Figure 3D) and C3a (p=0.007; Figure 3D), and higher C3a/C3 ratios (p=0.001; Figure 3D) compared to the WWH. MWH also had a greater decrease in C1-INH levels post-vaccine (p=0.003; Figure 3D), supporting the hypothesis that there is less inhibition of the classical pathway cascade in vaccinated MWH. Interestingly, there was not a significant change in C5a levels in either MWH or WWH post-vaccine (p=0.636; Figure 3D). Collectively, these data confirm that there is complement activation following vaccination in MWH that is absent in WWH, and that the cascade terminates post C3 deposition.

## Discussion

We show that the SARS-CoV-2 mRNA vaccines elicit an antibody response in WWH and MWH of differing magnitudes. Our cohort of women had a large proportion of vaccine non-responders that were present in both WWH and WWOH. Others have found a similar result, where approximately 50 percent of PLWH in their study were considered immunological non-responders, and this phenomena was not associated with CD4^+^ T cell counts [13]. This is especially interesting considering the majority of WWH in our study have CD4^+^ T cell counts considered to be in the healthy range (Table 1). Differences in the hormonal blood chemistry of the women in our study may be involved in the insufficient antibody response in our cohort [14, 15]. Most women in our cohort were over the age of 51 years, the median age of menopause in the general population. It has been shown that post-menopausal women specifically mount lower humoral responses to the influenza vaccine than younger women who have not undergone menopause [16], and that HIV infection increases this age-associated discrepancy in antibody response [15]. It has also been shown that age plays a significant role in antibody responses between men and women [17]. However, age did not seem to determine vaccine non-response (data not shown), indicating that age alone is not fully explanatory of the suboptimal antibody responses. Additionally, the timing of the initiation of antiretroviral therapy (ART) could be a factor [18].

We found that the complement system was also not significantly activated in vaccinated WWH. This finding contrasts with the complement activation we observed post-vaccine in MWH [3]. When investigating the mechanisms of complement cascade regulation, we found that a decline in C1-INH could be the driving force behind the increase in complement activity in MWH. Although we only measured anti-S1 ADCA, the lower levels of C1-IHN implies that ADCA should be elevated for all antigens in MWH. Indeed, a recent study postulates that complement activation in general could be a mechanism of virus neutralization enhancement that improves the potency of neutralizing antibodies [19] and antigen opsonization. This mechanism could contribute to the overall antibody-mediated protection to SARS-CoV-2 in MWH. This is in tandem with our finding of no change in C5a levels post-vaccine, indicating that the complement activation does not progress to a Membrane Attack Complex which could lead to a greater risk for comorbidities [3, 12, 20]. Importantly, we did not find that WWH had a significant change in C1-INH levels post-vaccine and lower antibody response when compared to the MWH. This adds to the greater picture of a more limited classical complement cascade activation in our cohort of women. Of note, our present study is limited by the small sample size and heterogenous nature of the two cohorts, restricting the generalizability of the findings outside our cohort.

Our study highlights a sex-based difference in the immune response to the SARS-CoV-2 mRNA vaccines and potentially to other vaccines in PWH. It also furthers the idea that reduced inhibition of the ADCA could be a compensatory mechanism for MWH to overcome the reduced antibody production. Our findings are pertinent in considering future SARS-CoV-2 immunization programs for women with and without HIV.

## Data Availability

All data produced in the present study are available upon reasonable request to the authors.

## Notes

## Acknowledgments

We thank the participants of the MACS/WIHS Combined Cohort Study for their outstanding support.

## Author contributions

ETAM, CRR, RBM, NSC, PT and DJT conceptualized and designed the study and wrote the protocol. DJT wrote the first draft of the manuscript and referred to appropriate literature and is the main author responsible for formal data analysis. DJT, ETAM, RBM, NSC, CRR and PT revised the manuscript content and reviewed and edited the manuscript. DJT performed all the serology tests. DJT performed the complement assays. DJT performed the data analysis. All authors agreed with and approved the final version of the manuscript.

## Data sharing

Deidentified patient data and a data dictionary are available following publication upon request to the corresponding author. Proposals will be reviewed and approved by the investigators and collaborators based on scientific merit. After approval of a proposal, data can be shared through a secure online platform after signing a data access agreement.

## References

1. Antinori A, Cicalini S, Meschi S, et al. Humoral and Cellular Immune Response Elicited by mRNA Vaccination Against Severe Acute Respiratory Syndrome Coronavirus 2 (SARS-CoV-2) in People Living With Human Immunodeficiency Virus Receiving Antiretroviral Therapy Based on Current CD4 T-Lymphocyte Count. Clinical Infectious Diseases 2022; 75:e552–e63.

2. Jedicke N, Stankov MV, Cossmann A, et al. Humoral immune response following prime and boost BNT162b2 vaccination in people living with HIV on antiretroviral therapy. HIV Medicine 2022; 23:558–63.

3. Tuttle DJ, Castanha PMS, Nasser A, et al. SARS-CoV-2 mRNA Vaccines Induce Greater Complement Activation and Decreased Viremia and Nef Antibodies in Men With HIV-1. The Journal of Infectious Diseases 2023; 229:1147–57.

4. Anticoli S, Capanna S, Volpin A, et al. Sex differences in response to HBV vaccination in a cohort of health care workers. Vaccine: X 2025; 22:100605.

5. Zhou S, Wang J, Lv M, et al. Long-term antibody trajectories after PPSV23 in elderly: Results from a 4-year follow-up study. Vaccine 2025; 48:126737.

6. Kurtovic L, Beeson JG. Complement Factors in COVID-19 Therapeutics and Vaccines. Trends in Immunology 2021; 42:94–103.

7. Bosmann M. Complement control for COVID-19. Sci Immunol 2021; 6.

8. Castanha PMS, Tuttle DJ, Kitsios GD, et al. IgG response to SARS-CoV-2 and seasonal coronaviruses contributes to complement overactivation in severe COVID-19 patients. The Journal of Infectious Diseases 2022.

9. Henry BM, Szergyuk I, de Oliveira MHS, et al. Complement levels at admission as a reflection of coronavirus disease 2019 (COVID-19) severity state. Journal of Medical Virology 2021; 93:5515–22.

10. Skendros P, Mitsios A, Chrysanthopoulou A, et al. Complement and tissue factor-enriched neutrophil extracellular traps are key drivers in COVID-19 immunothrombosis. J Clin Invest 2020; 130:6151–7.

11. D’Souza G, Bhondoekhan F, Benning L, et al. Characteristics of the MACS/WIHS Combined Cohort Study: Opportunities for Research on Aging With HIV in the Longest US Observational Study of HIV. American journal of epidemiology 2021; 190:1457–75.

12. Castanha PMS, Tuttle DJ, Kitsios GD, et al. Contribution of Coronavirus-Specific Immunoglobulin G Responses to Complement Overactivation in Patients with Severe Coronavirus Disease 2019 The Journal of Infectious Diseases 2022; 226:766–77.

13. Sisteré-Oró M, Andrade N, Wortmann DDJ, et al. Anti-SARS-COV-2 specific immunity in HIV immunological non-responders after mRNA-based COVID-19 vaccination. Frontiers in Immunology 2022; Volume 13-2022.

14. Parmigiani A, Alcaide ML, Freguja R, et al. Impaired Antibody Response to Influenza Vaccine in HIV-Infected and Uninfected Aging Women Is Associated with Immune Activation and Inflammation. PLOS ONE 2013; 8:e79816.

15. George VK, Pallikkuth S, Parmigiani A, et al. HIV infection Worsens Age-Associated Defects in Antibody Responses to Influenza Vaccine. The Journal of Infectious Diseases 2015; 211:1959–68.

16. Engelmann F, Rivera A, Park B, Messerle-Forbes M, Jensen JT, Messaoudi I. Impact of Estrogen Therapy on Lymphocyte Homeostasis and the Response to Seasonal Influenza Vaccine in Post-Menopausal Women. PLOS ONE 2016; 11:e0149045.

17. Klein SL, Marriott I, Fish EN. Sex-based differences in immune function and responses to vaccination. Transactions of The Royal Society of Tropical Medicine and Hygiene 2015; 109:9–15.

18. Moonsamy S, Suchard M, Madhi SA. Effect of HIV-exposure and timing of anti-retroviral treatment on immunogenicity of trivalent live-attenuated polio vaccine in infants. PLOS ONE 2019; 14:e0215079.

19. Mellors J, Dhaliwal R, Longet S, et al. Complement-mediated enhancement of SARS-CoV-2 antibody neutralisation potency in vaccinated individuals. Nature Communications 2025; 16:2666.

20. Taton M, Willems F, Widomski C, et al. HIV-related immune activation attenuates polyfunctional IgG and memory B-cell responses to Tdap immunization during pregnancy. eBioMedicine 2024; 104:105179.

