## Supplemental Methods and Figure 1 for "Sex, age and HIV-1 status influence complement activation against SARS-CoV-2 mRNA vaccine Spike protein"

**SUPPLEMENTAL MEHTODS**

*Complement activation assay.* Complement activation was assessed using a bead-based flow cytometry assay. SARS-CoV-2 WA S1 (SinoBiological, cat# 40591-V08H) were biotinylated using a Sulfo-NHS-LC biotin kit (Thermo Scientific, cat# 21935), and incubated with red 1.0µm neutravidin beads at a 1ug:1uL ratio (Thermo Fisher, cat# F8775) for 2h at 37°C. The beads were washed twice with a 5% BSA-PBS solution and stored at 4°C in a 0.1% BSA-PBS solution until use. Serum samples were diluted 1:10 in 0.1% BSA-PBS, added to a 96-well U bottom plate and heat inactivated at 56°C for 30 minutes. 10µL of bead solution were added to each well, and the plate was incubated at 37°C for 2 h. After incubation, guinea pig serum (MP Biomedicals, cat# 0855845) diluted 1:50 was added to each well and incubated at 37°C for 15 minutes. The plate was centrifuged at 2000g for 5 minutes, decanted, washed with 15mM EDTA, and centrifuged as described above. A FITC-conjugated α-C3 monoclonal antibody (MP Biomedicals, cat# 0855385), diluted 1:100 in PBS was added and incubated at room temperature in the dark for 15 minutes. The plate was washed twice with PBS and centrifuged as described above. The beads were fixed with 4% PFA for 20 minutes and resuspended in 100µL of PBS. The beads were then assessed on a BD FACSymphony. Each experiment was independently performed three separate times, and one run was chosen as a representative of these three.

*ELISA assays.* Briefly, recombinant spike S1 and N proteins (2µg/ml), or antibodies against C3, C3a, C5a or CFH were used to coat high-binding, 96-well polystyrene plates (Corning) overnight at 4°C in 0.2M carbonate/bicarbonate buffer in a humidified chamber. A goat Fab_2_ α-human IgG [H+L]-HRP secondary antibody (Jackson Immunoresearch; cat# 109-036-003) was used to quantify total IgG, and biotin antibodies in combination with a streptavidin-HRP conjugate was used to detect complement factors. Optical densities at a wavelength of 450nm (OD450nm) were measured using SpectraMax Plus PC380 microplate spectrophotometer using SoftMax Pro software version 6.4 (Molecular Devices, USA). Cut-off values were calculated as the mean OD450nm of the negative control plus 3 times the standard deviation. Endpoint antibody titers were calculated using 4-parameter non-linear regression on GraphPad Prism 10.1. Samples were run in duplicate, and the measurements were averaged.

**SUPPLEMENTAL FIGURES**

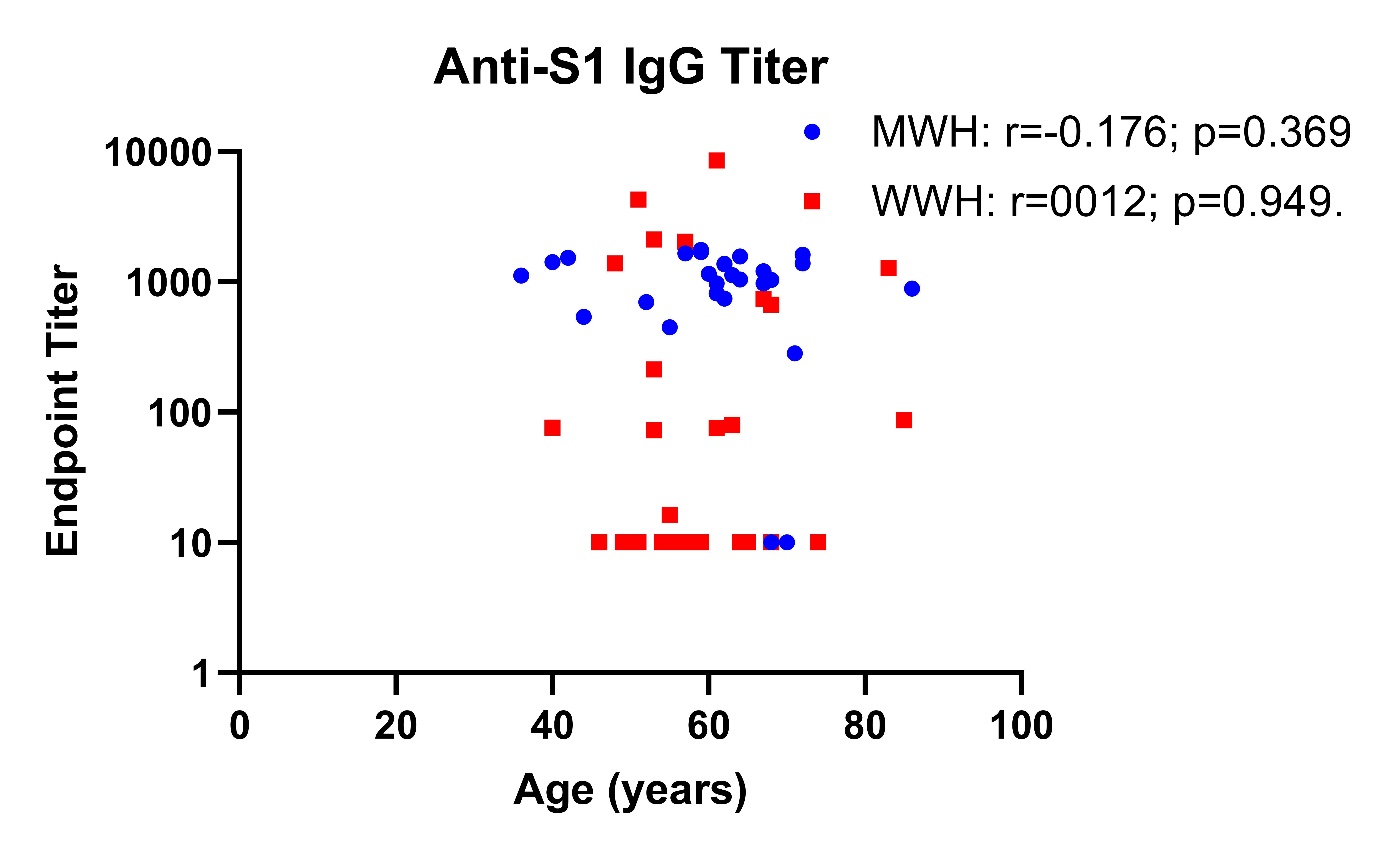

**Figure S1. Anti-S1 IgG titer by age for MWH and WWH.** Anti-S1 IgG levels in WWH and MWH were measured by indirect ELISA. Participants are anti-N IgG negative, and two-dose vaccinated with a SARS-CoV-2 mRNA vaccine. Spearman correlation was used to compare both variables. A p-value of 0.05 is considered statistically significant. Abbreviations: IgG, immunoglobulin G; WWH, women with HIV-1; MWH, men with HIV-1; N, nucleocapsid.
